# Performance of Thrombectomy-Capable, Comprehensive, and Primary Stroke Centers in Reperfusion Therapies for Acute Ischemic Stroke: Report from the Get With The Guidelines Stroke Registry

**DOI:** 10.1101/2023.07.05.23292270

**Authors:** Radoslav Raychev, Jie-Lena Sun, Lee Schwamm, Eric E. Smith, Gregg C. Fonarow, Steven R. Messé, Ying Xian, Karen Chiswell, Rosalia Blanco, Brian Mac Grory, Jeffrey L. Saver

## Abstract

**Background:** The thrombectomy-capable stroke center (TSC) is a recently introduced intermediate tier of accreditation for hospitals caring for patients with acute ischemic stroke (AIS). The comparative quality and clinical outcomes of reperfusion therapies at TSCs, primary stroke centers (PSCs), and comprehensive stroke centers (CSCs) has not been well delineated.

**Methods:** We conducted a retrospective, observational, cohort study from 2018-2020 that included patients with AIS who received endovascular (EVT) and/or intravenous (IVT) reperfusion therapies at CSC, TSC, or PSC. Participants were recruited from Get With The Guidelines–Stroke registry. Study endpoints included timeliness of IVT and EVT, successful reperfusion, discharge destination, discharge mortality, and functional independence at discharge.

**Results:** Among 84,903 included patients, 48,682 received EVT, of whom 73% were treated at CSCs, 22% at PSCs, and 4% at TSCs. The median annual EVT volume was 76 for CSCs, 55 for TSCs, and 32 for PSCs. Patient differences by center status included higher NIHSS, longer onset-to-arrival time, and higher transfer-in rates for CSC/TSC/PSC, respectively. In adjusted analyses, the likelihood of achieving the goal door-to-needle time was higher in CSCs compared to PSCs (OR 1.39; 95% CI 1.17-1.66) and in TSCs compared to PSCs (OR 1.45; 95% CI 1.08-1.96). Similarly, the odds of achieving the goal door-to-puncture time were higher in CSCs compared to PSCs (OR 1.58; 95% CI 1.13-2.21). CSCs and TSCs also demonstrated better clinical efficacy outcomes compared to PSCs. The odds of discharge to home or rehabilitation were higher in CSCs compared to PSCs (OR 1.18; 95% CI 1.06-1.31), while the odds of in-hospital mortality/discharge to hospice were lower in both CSCs compared to PSCs (OR 0.87; 95% CI 0.81-0.94) and TSCs compared to PSCs (OR 0.86; 95% CI 0.75-0.98). There were no significant differences in any of the quality-of-care metrics and clinical outcomes between TSCs and CSCs.

**Conclusions:** In this study representing national US practice, CSCs and TSCs exceeded PSCs in key quality-of-care reperfusion metrics and outcomes, whereas TSCs and CSCs demonstrated similar performance. Considering that over one-fifth of all EVT procedures during the study period were conducted at PSCs, it may be desirable to explore national initiatives aimed at facilitating the elevation of eligible PSCs to a higher certification status.

## INTRODUCTION

Endovascular therapy (EVT) for treatment of acute ischemic stroke (AIS) associated large vessel occlusion (LVO) increased substantially in the US following results of pivotal trials in 2015^1^, prompting reorganization of stroke systems of care. Despite the widespread surge in EVT volumes, regional direct access to comprehensive stroke centers (CSC) offering advanced endovascular capabilities remains limited. A recent study demonstrated that only one-third of the US population has adequate timely direct access to EVT.^2^ As a result, EVT access for many patients requires interfacility transfer, incurring delays of care with negative impacts on clinical outcome.^3, 4^ Increasing the overall number of hospitals performing high quality EVT and routing patients directly to those centers could facilitate widespread direct access to thrombectomy and improve clinical outcomes.

For this reason, in 2018, hospital accrediting bodies introduced a new, intermediate tier for hospital certification - Thrombectomy-Capable Stroke Centers (TSCs). A program certified as a TSC is required to maintain EVT-specific volumes and capabilities, including 24/7 neurointerventional team, 24/7 neuro ICU care for stroke patients, advanced neuroimaging, and ability to collect and review data pertinent to reperfusion therapy (eTable 1). TSCs are also required to collect 5 comprehensive stroke quality measures (CSTK), addressing key performance metrics for patients who receive EVT, intravenous thrombolysis, or a combination of these therapies. Compared with PSCs, which receive certification to provide intravenous thrombolytic therapy and organized inpatient care, TSCs receive additional certification for provision of guideline adherent EVT. Compared with CSCs, TSCs are not certified for provision of complex treatments for other forms of cerebrovascular disease, including endovascular and open surgical treatment of cerebral aneurysms and arteriovenous malformations. A TSC certification program was launched by the Joint Commission (TJC) in collaboration with the American Heart Association/American Stroke Association in January 2018, followed by similar certification programs by all US certification agencies: Det Norske Veritas (DNV) - “Primary Plus”, Healthcare Facilities Accreditation Program (HFAP) - “Thrombectomy Stroke”, and Center for Improvement in Healthcare Quality (CIHQ) - “Thrombectomy-Capable”.

In the era before the introduction of TSCs, a comparison between CSCs and PSCs in key GWTG-Stroke in-hospital metrics demonstrated similar overall quality of care, with the exception of faster delivery of thrombolytic therapy in CSCs and lower risk-adjusted in-hospital mortality in PSCs.^5^ However, comparison of EVT-specific quality of care metrics in the current system of acute stroke care, including the newly established TSC-level certification, has not yet been examined. The goal of this study was to compare the quality of acute ischemic stroke and clinical outcome between PSCs, TSCs, and CSCs within the GWTG-Stroke registry

## METHODS

### Study Setting

To ensure uniform adherence to evidence-based practices and stroke treatment guidelines, certified stroke centers participate in the GWTG-Stroke Registry. GWTG-Stroke was developed by the American Heart Association (AHA) to improve the quality of care for patients hospitalized with stroke. The program uses an online, interactive assessment tool to improve stroke care through increasing compliance with published guideline recommendations. Participation in the GWTG-Stroke registry has been shown to be associated with higher uptake of evidenced-base acute stroke practices and improved clinical outcomes. ^6, 7 8^

We performed a retrospective, observational, cohort study based on the American Heart Association Get With the Guidelines–Stroke registry, a voluntary, national stroke registry and performance improvement program with more than 6 million stroke admissions reported. The details and validity of the program have been described previously. ^9 10^ IQVIA is the data coordination center. The Duke Clinical Research Institute is the statistical coordinating center and analyzes deidentified data under an institutional review board–approved protocol. The requirement for obtaining informed consent was waived by local institutional review boards. This study followed the Strengthening the Reporting of Observational Studies in Epidemiology (STROBE) reporting guidelines.

### Study population

We analyzed all patients with ischemic stroke admitted from January 2018 to December 2020 who received acute reperfusion therapies in GWTG-Stroke hospitals certified as either PSCs, TSCs, or CSCs. The reperfusion therapies included EVT alone (EVT at this hospital), IVT alone (IVT at this hospital), or combined EVT (at this hospital) + IVT (at this hospital or outside hospital for patients with EVT at this hospital). Hospitals with less than 5 EVT cases per year were excluded. Patients with > 25% missing sex or pertinent history fields, in-hospital stroke onset, transferred-out to acute care facility, and missing NIHSS were also excluded (eFigure 1). Certification status was derived from publicly available Joint Commission (www.qualitycheck.org) and Det Norske Veritas (DNV) (https://www.dnvhealthcareportal.com) database. We did not verify certification status by HFAP (Thrombectomy Stroke), and CIHQ (Thrombectomy-Capable), and state agencies and hence did not include these certification assignations in this analysis.

**Figure 1.**
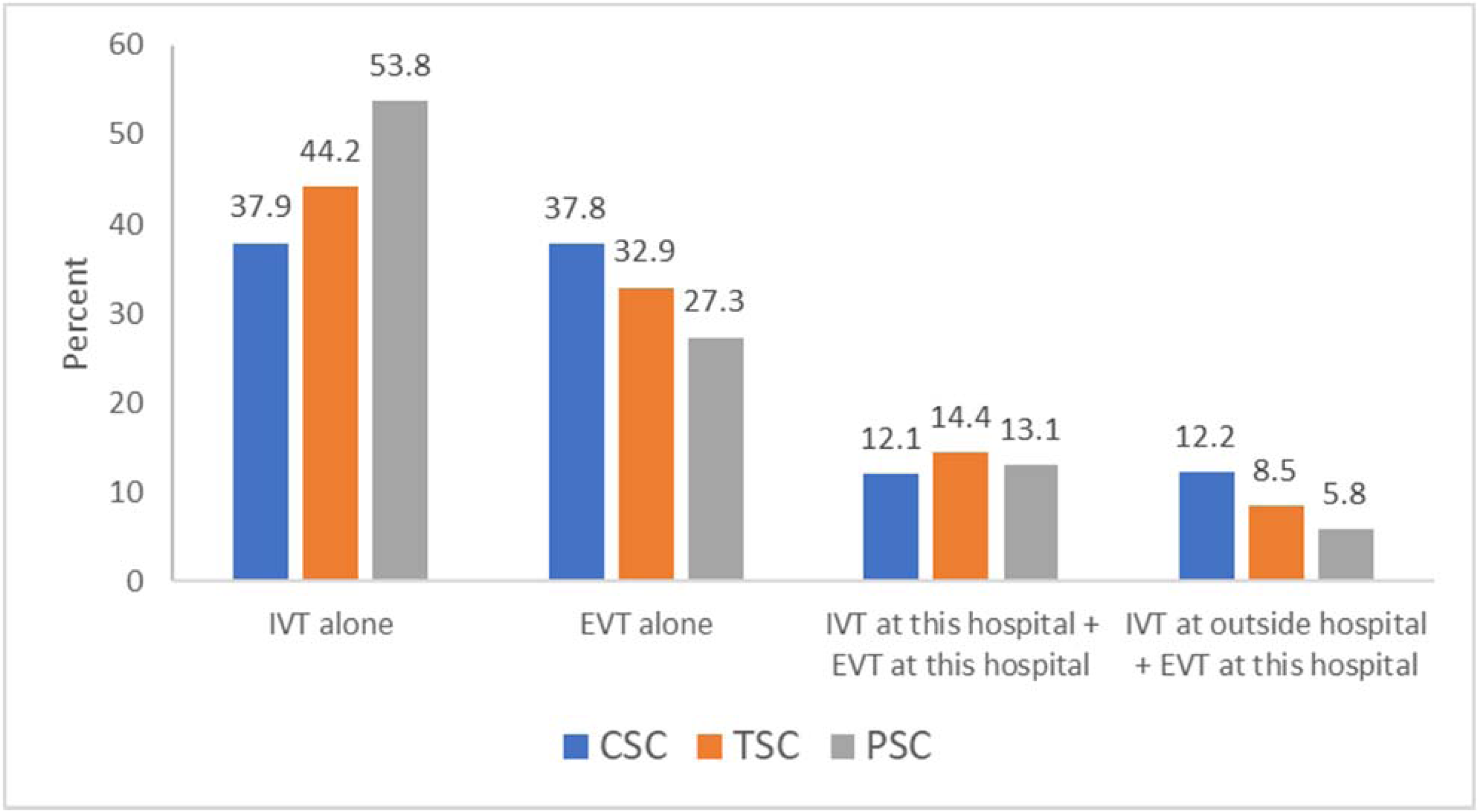
Distribution of reperfusion therapy per center.

### Characteristics of the study population

A total of 26 patient-level characteristics were included in the analyses: age, sex, race/ethnicity, health insurance, medical comorbidities (atrial fibrillation, previous stroke/TIA, CAD/prior MI, carotid artery stenosis, DM, PVD, hypertension, smoker, dyslipidemia, heart failure, renal insufficiency, sleep apnea, depression), medications (antihypertensive, lipid lowering, diabetic, antiplatelet, anticoagulation), National Institutes of Health Stroke Scale (NIHSS) score on arrival, mode of arrival, arrival during off hours, and onset-to-arrival time. In addition, 8 hospital characteristics were evaluated: stroke certification status, annual volume of ischemic stroke admissions, annual volume of IVT, annual volume of EVT, academic status, rural location, geographic region, and number of beds.

### Outcome measures

Process measure outcomes included timeliness of revascularization (door-to-needle time, door-to-puncture time, door-to-device time). The technical efficacy outcome measure was successful reperfusion, defined as modified thrombolysis in cerebral infarction (mTICI) score of 2b-3. The primary clinical efficacy outcome measure was functional independence (mRS 0-2) at discharge. Secondary clinical efficacy outcome measures were: mRS ordinal distribution on discharge; discharge to home; discharge to home or acute rehabilitation facility; ability to ambulate independently or with assistance at discharge. Safety outcome measures were in-hospital mortality; combined in-hospital mortality and discharge to hospice; and symptomatic intracranial hemorrhage.

### Statistical Analyses

We conducted univariate analyses comparing baseline characteristics (demographics, clinical variables, hospital characteristics), process metrics, technical outcomes, clinical efficacy outcomes, and safety outcomes by center status (PSC, TSC, CSC). The Kruskal-Wallis test was used for continuous variables and Pearson Chi-square or Fisher’s exact for categorical variables. Counts and percentages are presented for categorical/binary variables, and median (25^th^, 75^th^) and Mean (SD) for continuous variables.

Multivariable logistic regression models were generated to examine the association between the pre-defined process of care and clinical outcomes and stroke center certification status. We used generalized estimating equations (GEE) to account for hospital clustering of patients. The adjusted models were controlled for the multiple patient and hospital-level characteristics detailed above. All continuous variables included in the models were evaluated for nonlinearity with the outcome using restricted cubic splines and the likelihood ratio test, and splines were used for those that violated the linearity assumption. SAS (version 9.4; SAS Institute Inc.) software was used for all statistical analyses. All *P* values were 2-sided and statistical significance was defined as a *P* value of less than .05 for models.

Variables with missing data were not imputed for univariate tables. Patient characteristics with < 25% missing data were imputed before entering the model. By consensus, patient medical history or medication prior to admission missing values were imputed to “No” (missing interpreted as absence of history or medications).

## RESULTS

Application of study entry criteria yielded a total of 84,903 patients from 383 sites for analysis (eTable 1), including 57,306 (67.5%) at CSCs, 3579 (4.2%) at TSCs, and 24,018 (28.3%) at PSCs. During the study period, the number of TSCs increased from 7 in 2018 to 31 in 2019 and 35 in 2020. By the end of the study period, 185 were CSCs, 35 TSCs, and 163 PSCs. Reperfusion therapies were EVT alone in 29,373 (34.6%), EVT+IVT at the EVT-performing site in 10,618 (12.5%), EVT at the EVT-performing site and IVT at an outside hospital in 8,691 (10.2%), and IVT alone (at this site) in 36,221 (42.7%). Procedural case mix is shown in Figure 1. The proportion of cases treated with EVT alone was highest at CSCs, followed by TSCs, then PSCs. Similarly, the proportion of patients treated with IVT at an outside hospital followed by EVT at the EVT-performing site hospital was also highest at CSCs, followed by TSCs, then PSCs.

Conversely, the proportion of cases treated with IVT alone was highest at PSCs, followed by TSCs, then CSCs. The distribution of case volumes at CSCs, TSCs, and PSCs is shown in Figure 2. Annual EVT volumes were highest at CSCs (median 76), followed by TSCs (median 55), and PSCs (median 32).

**Figure 2.**
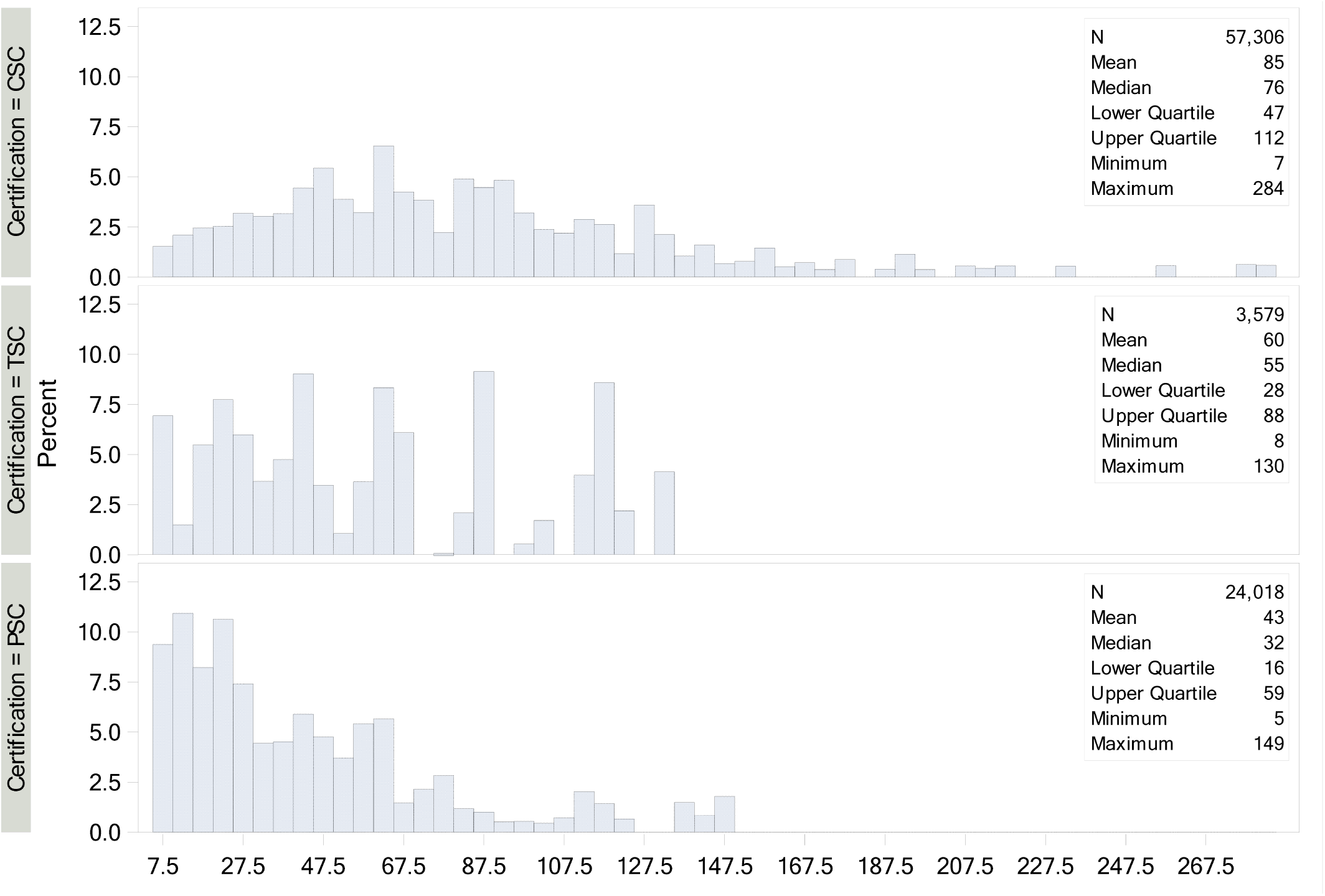
Distribution of EVT volume per center.

Differences in patient-level and hospital-level baseline characteristics by center status are summarized in Table 1. Multiple differences in baseline characteristics per center status were identified, including higher NHISS (12 vs 11 vs 10), longer onset-to-arrival time (2.3h vs 1.7h vs 1.5h), and higher transfer rates (33.4% vs 23.1% vs 16.6%) at CSCs vs TSCs vs PSCs. The overall median annual volume of ischemic stroke admissions was highest in CSCs (445) and equivalent in TSCs (286) and PSCs (295). The overall median annual volume of EVT was highest at CSCs (76.0) intermediate at TSCs (55) and lowest at PSCs (32). However, the proportion of all admitted ischemic stroke patients undergoing EVT was high at both TSCs (19.2%) and CSCs (17.0%), and lower at PSCs (10.8%).

**Table 1.** Patient and hospital characteristics at CSCs, TSCs, and PSCs

| Patient Characteristics | CSC<br>Patients n=57,306,<br>Sites=185 | TSC<br>Patients n=3579,<br>Sites=29 | PSC<br>Patients n=24018,<br>Sites=169 | P-Value |
| --- | --- | --- | --- | --- |
| Age, median (IQR) | 70 (59-81) | 72 (61-81) | 71 (59-80) | 0.0008 |
| Sex, female | 49.3% | 49.7% | 48.7% | 0.20 |
| Race-ethnicity |  |  |  |  |
| White, non-Hispanic | 64.6% | 65.7% | 67.6% | <.0001 |
| Black | 18.5% | 19.8% | 14.6% |  |
| Hispanic | 7.9% | 6.1% | 10.4% |  |
| Asian | 3.2% | 2.7% | 2.8% |  |
| Other | 5.9% | 5.6% | 4.6% |  |
| NIHSS | 12.0 (6.0-19.0) | 11.0 (5.0-18.0) | 10.0 (5.0-17.0) | <.0001 |
| Medical Hx |  |  |  |  |
| Hypertension | 72.6% | 76.0% | 73.0% | <.0001 |
| Dyslipidemia | 46.0% | 52.6% | 45.7% |  |
| Diabetes mellitus | 28.2% | 26.9% | 29.3% |  |
| Carotid stenosis | 2.9% | 3.3% | 2.9% |  |
| CAD/prior MI | 21.8% | 21.2% | 22.6% |  |
| Atrial fibrillation | 24.6% | 23.6% | 21.4% |  |
| Tobacco use | 19.1% | 16.8% | 18.2% |  |
| Prior stroke/TIA | 23.2% | 26.8% | 25.1% |  |
| Arrival Mode |  |  |  |  |
| EMS from home/scene | 56.1% | 64.3% | 66.7% | <.0001 |
| Private transport | 9.5% | 12.3% | 15.8% |  |
| Interfacility transfer | 33.4% | 23.1% | 16.6% |  |
| Mobile Stroke Unit | 0.6% | 0.2% | 0.4% |  |
| Arrival On-Hrs (v Off-Hrs) | 54.6% | 52.8% | 52.8% |  |
| LKW to arrival (hrs) | 2.3 (1.0-5.2) | 1.7 (0.8-4.0) | 1.5 (0.8-3.3) |  |
| Health Insurance |  |  |  |  |
| Private/VA/Other | 42.6% | 47.3% | 45.2% |  |
| Medicaid | 11.9% | 11.2% | 12.4% | <.0001 |
| Medicare | 40.5% | 38.5% | 36.4% |  |
| Self-Pay/No Insurance | 5.1% | 2.9% | 6.0% |  |
| ***Hospital Characteristics*** |  |  |  |  |
| Annual Volume of Ischemic Stroke Admissions (median) | 445 | 286 | 295 | <.0001 |
| Annual Volume of IV Lytics (median) | 61.0 | 56.0 | 56.0 |  |
| Proportion of ischemic stroke patients treated with IVT | 13.7% | 19.6% | 18.9% |  |
| Annual Volume of EVTs (median) | 76.0 | 55.0 | 32.0 |  |
| Proportion of ischemic stroke patients treated with EVT | 17.0% | 19.2% | 10.8% |  |
| Hospital size, # of beds (median) | 445 | 417 | 417 |  |
| Academic hospital | 95.7% | 81.2% | 82.0% |  |
| Region |  |  |  |  |
| Northeast | 23.6% | 4.3% | 10.2% | <.0001 |
| Midwest | 23.0% | 22.8% | 15.8% |  |
| South | 38.7% | 44.8% | 49.1% |  |
| West | 14.7% | 28.1% | 24.9% |  |
| Teaching Hospital | 95.7% | 81.2% | 82.0% |  |
| Rural Location | 0.0% | 0.0% | 1.9% |  |

Process measure performance for reperfusion therapy by center type is summarized in Table 2. Overall, CSCs and TSCs performed similarly in all time to treatment metrics; PSCs had slightly worse performance. The median door-to-needle time in PSCs was 7 min slower than that at TSCs and 5 min slower than that at CSCs. The median door-to-puncture time in direct-arriving patients in PSCs was 12 min slower than that at TSCs and 16 min slower than that at CSCs.

**Table 2.** Reperfusion process metrics

| Metric | CSC | TSC | PSC |
| --- | --- | --- | --- |
| Door-to-Needle (min) | 42 (30-59) | 40 (28-57) | 47 (34-64) |
| Door to Needle ≤ 45 min | 76.8% | 78.2% | 70.8% |
| Door to Needle ≤ 60 min | 77.2% | 78.0% | 71.1% |
| Door to puncture– All (min) | 74 (44-105) | 79 (47-113) | 94 (62-131) |
| Door to puncture within target | 39.1% | 39.0% | 26.7% |
| Direct arriving patients, door to puncture ≤ 75 min | 30.5% | 29.0% | 17.2% |
| Door to puncture time ≤ 45 min (transfer) | 47.3% | 55.3% | 43.2% |
| Door to first pass time – All (min) | 96 (64-132) | 103 (71-141) | 115 (81-157) |
| Direct-arriving patients | 115 (90-149) | 119 (93-151) | 131 (102-173) |
| Interfacility transfer patients | 71 (47-106) | 66 (43-102) | 75 (44-117) |
| Door to first pass within target | 32.5% | 31.0% | 23.9% |
| Door to first pass ≤ 90 min (direct) | 25.1% | 22.7% | 15.5% |
| Door to first pass ≤ 60 min (transfer) | 39.6% | 44.2% | 38.7% |
| Successful reperfusion (mTICI 2b-3) | 89.4% | 90.0% | 87.2% |

Successful reperfusion (mTICI 2b-3) rates were highest in TSCs (90%) and CSCs (89.4%), and mildly lower in PSCs (87.2%). Clinical outcomes by center type are shown in Table 3. For efficacy outcomes, discharge to home or acute rehabilitation was equally frequent at all facility types, ambulation with or without assistance at discharge mildly lower at TSCs than CSCs and PSCs, and functional independence (mRS 0-2) at discharge mildly higher at PSCs than CSCs and TSCs. For safety outcomes, in-hospital death or discharge to hospice was equally frequent at all facility types while symptomatic intracranial hemorrhage was mildly less frequent at TSCs than at CSCs and PSCs.

**Table 3.**
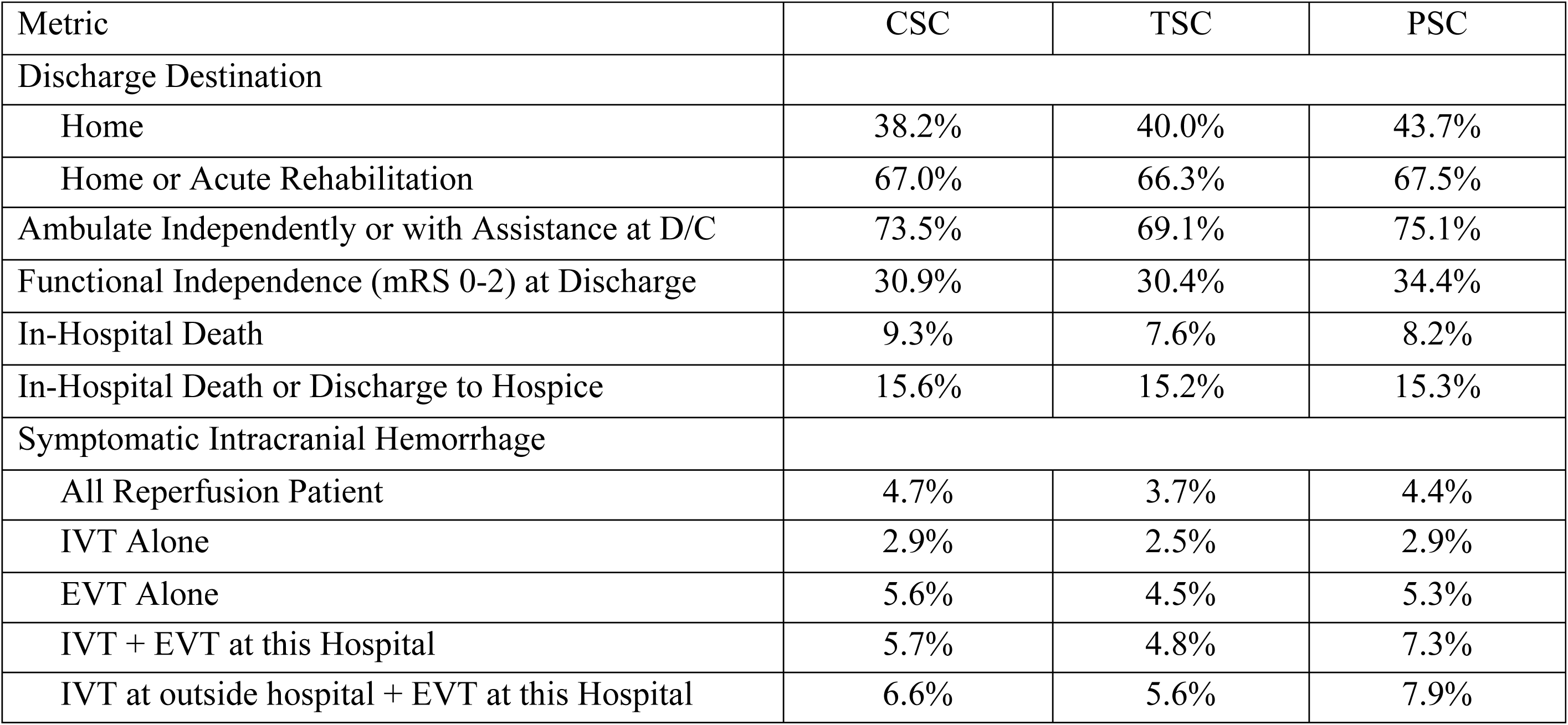
Clinical outcomes

Case mix-adjusted comparisons by center status for process metrics and clinical outcomes are shown in Table 4. The overall results highlight substantial differences in key quality of care metrics and clinical outcomes based on certification status. For speed of care and achievement of successful reperfusion process outcomes, comparative patterns paralleled those in unadjusted analysis, with CSCs and TSCs substantially exceeding PSCs. The likelihood of achieving the goal door-to-needle time was higher in CSCs compared to PSCs (OR 1.39; 95% CI 1.17-1.66) and in TSCs compared to PSCs (OR 1.45; 95% CI 1.08-1.96). Similarly, the odds of achieving the goal door-to-puncture time were higher in CSCs compared to PSCs (OR 1.58; 95% CI 1.13-2.21). CSCs and TSCs also demonstrated better clinical efficacy outcomes compared to PSCs. The odds of discharge to home or rehabilitation were higher in CSCs compared to PSCs (OR 1.18; 95% CI 1.06-1.31). Additionally, the odds of in-hospital mortality/discharge to hospice were lower in both CSCs compared to PSCs (OR 0.87; 95% CI 0.81-0.94) and TSCs compared to PSCs (OR 0.86; 95% CI 0.75-0.98). There were no significant differences in any of the quality-of-care metrics and clinical outcomes between TSCs and CSCs.

**Table 4.**
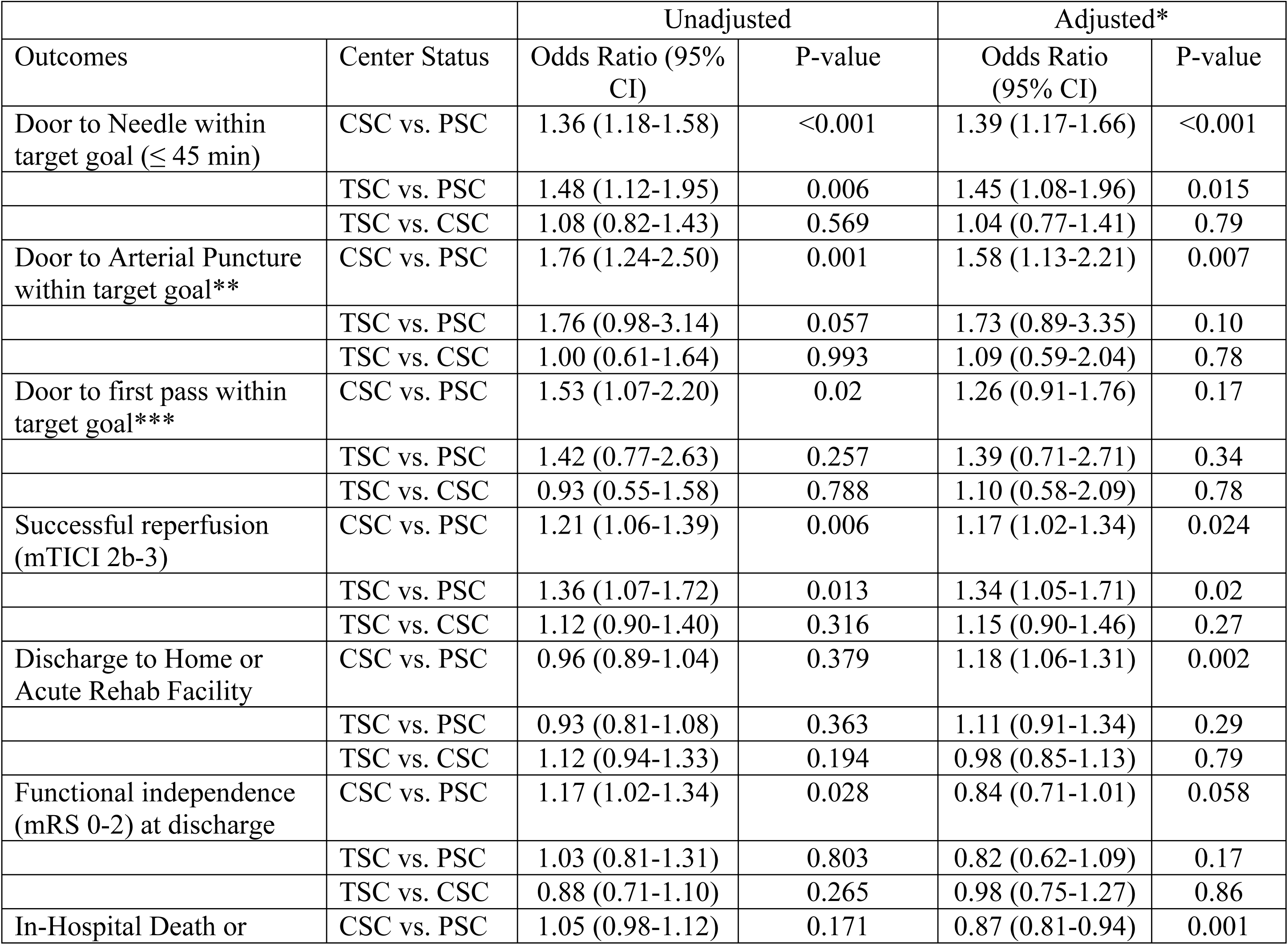
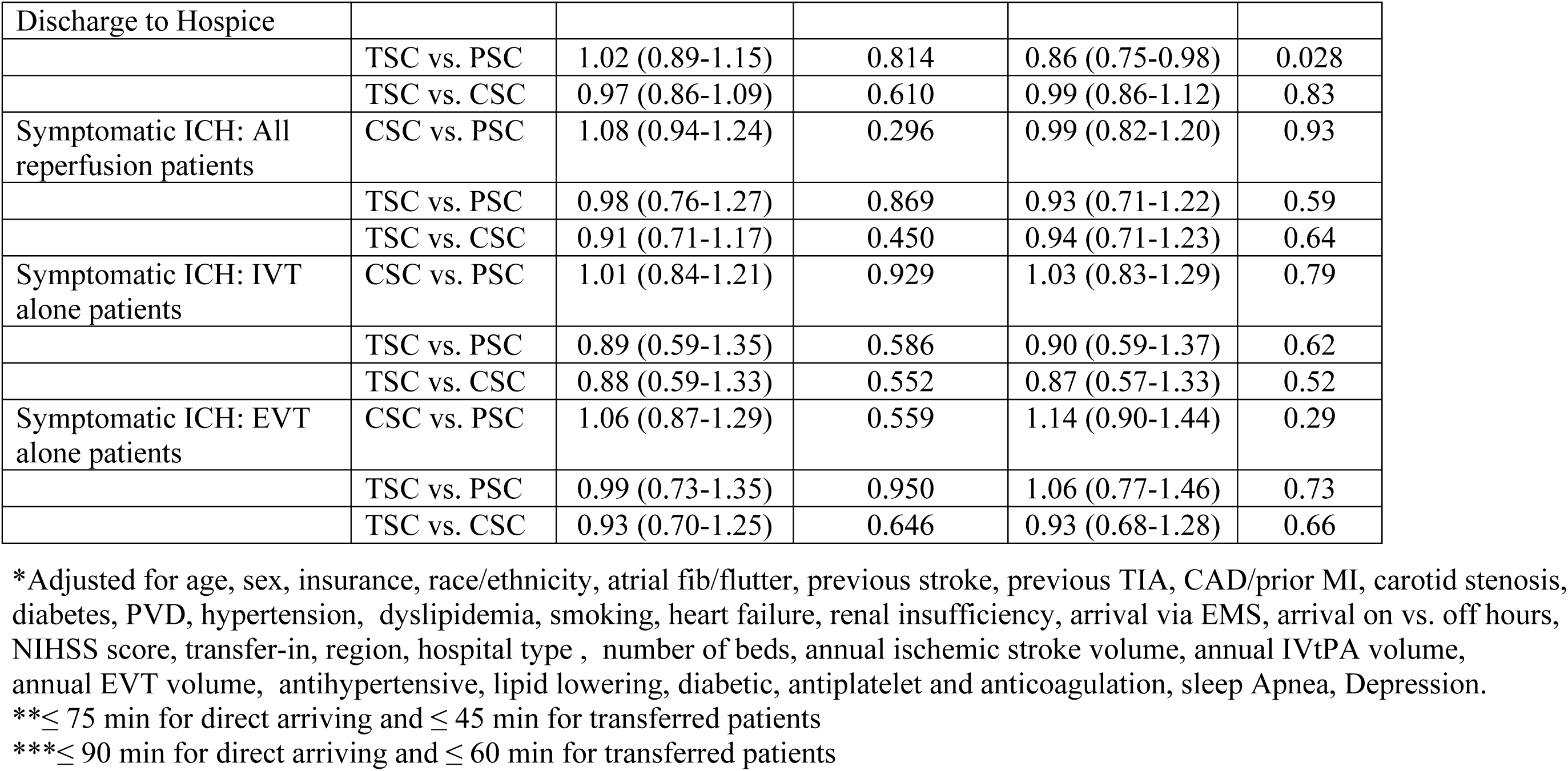
Comparisons of process and clinical outcomes by center status, unadjusted and adjusted.

## DISCUSSION

In this study of over 84,000 acute ischemic stroke patients treated with acute reperfusion therapy in routine clinical practice, similar high quality of care was achieved at CSCs and TSCs, and they were both superior to PSCs. Clinical outcomes at discharge were largely similar among all facility types, except for a lower rate of in-hospital mortality or discharge to hospice at CSCs and TSCs than at PSCs. TSCs accounted for a small but growing proportion of reperfusion-treated patients during the study period, and geographical variation was noted, with TSCs providing 1 in every 15 reperfusion therapies in the West, 1 in every 22 reperfusion therapies in the Midwest and South, and 1 in every 100 reperfusion therapies in the Northeast.

This study importantly extends the literature regarding the association of stroke center accreditation status and process and clinical outcomes. A prior analysis of GWTG-Stroke-participating hospitals compared performance of CSCs and PSCs, but was conducted in the pre-TSC era and analyzed all patients with ischemic stroke, not just those undergoing reperfusion therapy.^5^ Another study only analyzed the subset of CSCs and TSCs certified by one certifying organization, did not analyze PSCs, evaluated only EVT patients and not those receiving IVT, assessed only 2 process and 1 safety metric, and did not perform analyses adjusted for case mix.^11^

A particularly noteworthy aspect of our findings is the very similar performance of CSCs and TSCs on process, clinical efficacy, and safety metrics. These findings may help to allay concerns that the improved access to endovascular thrombectomy afforded by TSCs in regions with limited CSC availability would be offset by reduced quality of care at TSCs. The results support the concept of requiring facilities that perform EVT to track and report EVT-specific care quality metrics to independent certifying bodies, regardless of their stroke center status. Currently, only TSC and CSC, but not PSCs are required to report these important quality metrics, which promotes hospital and provider treatment commitment to a similar degree at both types of facilities certified for EVT care.

Our risk-adjusted findings of substantially faster care metrics, higher discharge to home or acute rehabilitation, and lower mortality or discharge to hospice at CSCs and TSCs vs PSCs in centers with advanced certification status can be explained by several factors. PSCs likely generally have less personnel and equipment resources for acute reperfusion care than CSCs and TSCs. PSCs do not have the incentive of annual certifying body review to drive continuous quality improvement for EVT. The substantially faster timeliness of IVT and EVT achieved in TSCs and CSCs likely contributed to the better clinical outcomes and re-affirm the well-established impact of faster revascularization on functional outcome in patients treated with intravenous and endovascular reperfusion therapy for AIS. ^12 13^ These results highlight the importance of rigorous implementation of evidence-based practices to meet the stringent requirements for TSCs and CSCs set by DNV and JC standards, including 24/7 availability of thrombectomy, neurology, and neurocritical care expertise, and multiple other rigorous competency standards for hospital providers and nursing staff. These requirements ensure a streamlined advanced approach to stroke care and ultimately translate into better outcomes for stroke patients. ^6 7, 8 5 14^

The higher revascularization rates achieved in TSCs and CSCs in comparison to PSCs is another important variable that likely contributed to better clinical outcomes. Successful revascularization is the most significant factor defining clinical outcome after EVT and is heavily influenced by operator’s experience. ^15^ CSCs and TSCs clearly outnumbered PSCs in annual EVT volume in our study. Multiple studies have demonstrated an association between higher annual EVT volumes and higher reperfusion rates with better clinical outcomes.^16 17 18^ It is important to highlight that median annual EVT volume in PSCs (32) in our study was twice as high as the minimal requirement (15) for obtaining more advanced certification status with some PSCs reaching up to 149 thrombectomies per year (Fig. 1). These results indicate that many of the PSCs participating in this study met annual volume criteria for more advanced certification status.

We also observed substantially higher EVT treatment rates proportional to annual stroke admission in CSCs and TSCs in comparison to PSCs. AIS due to LVO contributes disproportionally to stroke dependence and death and our real-practice data confirms the greater readiness of CSCs and TSCs than PSCs to deliver appropriate EVT treatment. ^19^ The higher EVT rates in CSCs and TSCs can be explained by implementation of processes specific to regional systems of care, including transfer protocols, EMS education initiatives, pre-hospital LVO recognition, preferential triage to advanced centers, and establishment of in-hospital protocols for streamlined workflow. Implementation of pre-hospital LVO recognition and preferential triage to CSC in regional systems of stroke care has been associated with increased EVT treatment rates in multiple simulation-based and real-world practice studies.^20 21 22 23^ An analysis of one regional acute stroke system of care in which EMS triaged all suspected LVOs to EMS-designated stroke-receiving centers that are required to be EVT-ready, regardless of their certification status (primary vs comprehensive) also demonstrated a substantial increase in reperfusion therapies with reduced in-hospital mortality over time.^24^ However, such a system of care may lead to dilution of procedural volumes from comprehensive stroke centers to centers with less advanced certification and less stringent process-of-care metrics with potential negative impact on clinical outcome as demonstrated by our data.

This study has limitations. First, we analyzed TSC performance at the start of the TSC era; as a result, findings are of a snapshot of time when this care model was first being introduced and only a relatively low number of TSCs were providing care. It is possible that TSC performance may improve further as time goes on, but it is also worth noting that participating TSCs were likely previously high-performing PSCs. This is evident in the superior performance of TSCs compared to PSCs on IVT quality metrics in addition to the EVT measures. Second, we used only DNV and TJC for determination of certification status as none of the other two main national accreditation agencies in the US (HFAP and CIHQ) had formally started the certification process for TSC during the study period. It is also very likely that some of the EVT-performing PSCs in our study were certified as CSCs or TSCs by local state agencies or were included in prehospital routing policies for suspected LVO patients. However, local state criteria for advanced certification status may be highly variable, while the DNV and TJC certification criteria are uniform across all geographical locations. Furthermore, these national certifying agencies have well-established protocols, ensuring close surveillance and strict adherence to GWTG metrics. Third is the site-reported nature of the analyzed data, which is dependent on the accuracy and completeness of abstraction from the medical record. Although the potential exists for selection bias, comparison of entered patients with national Medicare data sets has confirmed the representativeness of the GWTG-Stroke population. ^25^ Fourth, the clinical outcomes reported in this study are limited to short-term outcome with no data on long-term disability and functional outcome. However, other studies have shown that functional status at time of acute hospital discharge, including ambulatory status and discharge destination, correlates strongly with long-term global disability outcomes at 3 months.^26, 27^ Finally, residual measured and unmeasured confounding may have influenced some of these findings.

## CONCLUSIONS

Our study of national US practice reveals that CSCs and TSCs surpassed PSCs in essential reperfusion metrics and outcomes. TSCs demonstrated comparable performance to CSCs. Given the considerable number of EVT procedures performed at PSCs, there is a compelling rationale for exploring initiatives to support eligible PSCs in achieving higher certification status.

### Sources of funding

The Get With The Guidelines®–Stroke (GWTG-Stroke) program is provided by the American Heart Association/American Stroke Association. GWTG-Stroke is sponsored, in part, by Novartis, Boehringer Ingelheim and Eli Lilly Diabetes Alliance, Novo Nordisk, Sanofi, AstraZeneca, Bayer and Alexion Pharmaceuticals.

### Disclosures

RR reports equity for consulting for Rapid Medical, Perflow Medical, Spartan Micro, and fees for consulting for Boehringer Ingelheim, NeuroVasc, Medtronic

BMG reports funding from the National Heart, Lung, and Blood Institute (K23HL161426).

EES reports no relevant disclosures.

GCF reports receiving research support from the Patient Centered Outcome Research Institute and the National Institutes of Health, and employee of University of California which holds a patent on an endovascular device for stroke.

SRM reports receiving research support from the National Institutes of Health, WL Gore, Mallinkrodt, Biogen; consulting fees from Terumo, Boston Scientific, and EmStop; and reports equity in Neuralert Technologies

LS reports fees for consulting regarding trial design and conduct to Genentech (late window thrombolysis) and as a steering committee member (TIMELESS NCT03785678); user interface design and usability to LifeImage; stroke systems of care design to the Massachusetts Dept of Public Health; as a member of a Data Safety Monitoring Board (DSMB) for Penumbra (MIND NCT03342664) and for Diffusion Pharma PHAST-TSC NCT03763929); Serving as National PI for Medtronic (Stroke AF NCT02700945); Site PI, StrokeNet Network NINDS (New England Regional Coordinating Center U24NS107243) PRIME® Education: stroke systems of care and improving time to thrombolysis, and Boehringer-Ingelheim: stroke systems of care and improving time to thrombolysis.

## Data Availability

Data types: Deidentified participant data How to access data: Data were collected by the American Heart Association (the steward of the data according to contracts between the American Heart Association and participating hospitals) and are stored securely at the Duke Clinical Research Institute (DCRI). Given that data were collected for clinical care and quality improvement, rather than primarily for research, data sharing agreements require an application process for other researchers to access the data. Interested researchers can submit proposals to utilize Get With The Guidelines for research purposes, including for validation purposes. Proposals can be submitted at http://www.heart.org/qualityresearch. Additional information regarding the statistical analysis plan and analytic code may also be available from DCRI upon request. When available: With publication Who can access the data: Given that data were collected for clinical care and quality improvement, rather than primarily for research, data sharing agreements require an application process for other researchers to access the data. Interested researchers can submit proposals to utilize Get With The Guidelines for research purposes, including for validation purposes. Proposals can be submitted at http://www.heart.org/qualityresearch. Additional information regarding the statistical analysis plan and analytic code may also be available from DCRI upon request.

